# Symptoms of problematic alcohol use differ in their genetic associations with comorbid internalizing, externalizing, and neurodevelopmental psychiatric disorders

**DOI:** 10.1101/2025.10.20.25336759

**Authors:** Frances L. Wang, Dylan Maher, Daniel Bustamante, Kaitlin E. Bountress

**Author notes:** **Correspondence:** 3811 O’Hara St, Pittsburgh, PA 15213. Wang and Maher contributed equally to this work. **Declarations of competing interests:** none. **Primary Funding:** This study was funded by National Institutes of Health (NIH) grants K01 AA027757 (Wang), K01 AA028058 (Bountress), and R34 DA061267 (Bountress).

## Abstract

**Background and Aims:** Certain symptoms of problematic alcohol use (PAU) show associations with comorbid internalizing and externalizing disorders even after controlling for their common PAU factor. Outsized associations between PAU indicators and comorbid psychopathology may reflect distinct etiologic pathways or measurement characteristics that, if unaccounted for, could bias comorbidity estimates with PAU. Although these issues could represent a source of bias in estimates of genetic correlation with PAU, no studies have yet extended this work using genomic data.

**Design:** Genomic structural equation modeling and the Qtrait function identified PAU indicators that showed appreciable residual genetic correlations with eleven comorbid psychiatric disorders and whose associations did not operate strictly through the latent PAU factor.

**Setting:** Genome-wide association studies (GWAS) were conducted in a variety of international locations.

**Participants:** GWAS used in this study were conducted on 86,979 to 425,166 individuals of European ancestry.

**Measurements:** The primary measurements were GWAS summary statistics for various forms of internalizing, externalizing, and neurodevelopmental psychiatric disorders and nine indicators from the Alcohol Use Disorder Identification Test.

**Findings:** PAU indicators assessing alcohol-related consequences (i.e., *Injuries, Failed expectations, Guilt/Remorse*) each showed appreciable and positive residual genetic associations with multiple comorbid psychiatric conditions spanning various disorder spectra. *Alcohol Quantity*, *6+ Frequency*, *Blackouts*, and *Others concerned* did not show direct genetic relationships with comorbid disorders.

**Conclusions:** Alcohol-related consequences share unique genetic underpinnings with multiple psychiatric conditions apart from what is shared with their latent problematic alcohol use factor. Thus, alcohol-related consequences may unduly reflect dysfunction from comorbid psychiatric conditions or related third variables.

## INTRODUCTION

Problematic alcohol use (PAU) co-occurs at high rates with many psychiatric disorders across internalizing, externalizing, and psychotic spectra (1). Understanding the reasons for psychiatric comorbidity with PAU could elucidate novel shared risk mechanisms and transdiagnostic treatment targets. Research has shown that shared genetic factors contribute to the comorbidity between PAU and other psychiatric disorders, with moderate-to-high-genetic correlations reported with other substance use disorders, externalizing and neurodevelopmental disorders, internalizing disorders, and psychotic disorders (2).

Genetic correlations observed between PAU and other psychiatric disorders have been assumed to reflect meaningful shared genetic variance underlying these conditions (3,4). However, in these studies, PAU has commonly been defined as a sum of individual symptoms from dimensional scales like the Alcohol Use Disorders Identification Test (AUDIT; 5) or when thresholds for diagnostic criteria are surpassed (i.e., alcohol use disorder; AUD; 6). These common approaches to defining PAU assume that each symptom is equally informative in measuring it, which is likely not the case (7). Moreover, when these kinds of PAU-related constructs are studied in the context of psychiatric comorbidity, the assumption is that each PAU symptom is equally related to, and that no one PAU symptom has an outsized association with, each psychiatric condition (8). If this assumption is violated, it could bias genetic correlation estimates between PAU and other psychiatric disorders. Few studies to our knowledge have tested this important assumption.

The outsized influence of individual symptoms of PAU on certain psychiatric disorders has been previously theorized and empirically examined in the broader literature on AUD. Symptoms reflecting alcohol-related consequences (e.g., hazardous use; failure to fulfill role obligations; use despite social/interpersonal problems) may be especially susceptible to showing stronger-than-expected relationships with comorbid psychiatric conditions. Indeed, alcohol-related consequences may be difficult to attribute to alcohol pathology vs. third variables like comorbid psychiatric disorders (9). For example, research has shown an association between hazardous alcohol use (e.g., driving under the influence) and externalizing problems even after controlling for a latent AUD factor and alcohol consumption (10,11). Thus, hazardous alcohol use may reflect externalizing-related heedlessness in addition to alcohol-related compulsivity. Alcohol-related social/interpersonal problems and role interference also remained strongly tied to externalizing and internalizing problems after controlling for latent AUD and alcohol use (10–12). This link suggests that these PAU symptoms may pick up on functional impairment due to comorbid psychopathology in addition to alcohol pathology. Alternatively, findings could suggest that certain constellations of PAU symptoms are linked via unique genetic pathways to certain psychiatric conditions, which could inform treatment and prevention. However, no studies have examined whether certain indicators of PAU show especially strong genetic links with certain psychiatric disorders nor how accounting for them could influence estimates of genetic correlation between PAU and psychiatric disorders.

One prior investigation (13) conducted a genome-wide association study (GWAS) for each of the 10 items of the AUDIT, a widely used measure of PAU (5), and applied genomic structural equation modeling (SEM; 11) to investigate their latent genetic factor structure. They found that a two-factor model fit the data best. One factor contained three items pertaining to alcohol consumption and the second contained seven items pertaining to alcohol problems. The alcohol consumption and problems factors were highly correlated (*r*=0.80). As expected, these factors were positively correlated with alcohol dependence and psychiatric disorders (though more strongly for the alcohol problems factor) and negatively correlated with socioeconomic variables. In contrast, the residual variance of the item assessing frequency of alcohol use (i.e., the genetic variance in this item that was unrelated to all other AUDIT items) was *positively* correlated with socioeconomic status and *negatively* correlated with psychopathology. These findings served to reconcile prior work that showed similarly unexpected associations between alcohol consumption frequency and psychiatric/socioeconomic outcomes (15,16). The application of empirically derived weights to the AUDIT items in a genomic SEM framework in this study helped to reduce confounding or selection bias in measures of alcohol consumption and problems and identified the frequency-based measure of alcohol consumption as particularly susceptible to such biases.

The GWAS data on individual AUDIT items (13) permit a broader investigation of direct genetic relationships between AUDIT items and a range of psychiatric disorders after accounting for a common PAU factor(s) underlying AUDIT indicators. In this study, we first examined the dimensionality of the AUDIT indicators using parallel analysis and confirmatory factor analyses. Next, we examined whether individual symptoms of PAU had influential direct genetic associations with 11 psychiatric phenotypes after accounting for variation that was common to symptoms of PAU (i.e., their observed latent factor(s)). The psychiatric phenotypes examined included other substance use phenotypes (i.e., cannabis use disorder [CUD], opioid use disorder [OUD], cigarettes per day, drinks per week), externalizing/neurodevelopmental disorders (i.e., antisocial behavior [ASB], attention-deficit/hyperactivity disorder [ADHD]), internalizing disorders (i.e., post-traumatic stress disorder [PTSD], major depressive disorder [MDD]), compulsive disorders (i.e., obsessive compulsive disorder [OCD]), and psychotic disorders (i.e., schizophrenia [SCZ], bipolar disorder [BIP]). We also examined how accounting for specific direct genetic relationships between PAU indicators and other psychiatric conditions changed the genetic correlations between the PAU factor and each psychiatric trait. We hypothesized that AUDIT items assessing hazardous use, role interference, and social/interpersonal problems would show appreciable positive residual genetic associations (i.e., based on effect size thresholds, see methods), and that the frequency of alcohol use would show appreciable negative residual genetic associations, with various comorbid psychiatric conditions. Finally, we hypothesized that accounting for these associations would change the genetic correlations among PAU and these psychiatric conditions.

## METHOD

### Samples

We obtained summary statistics from GWAS of psychiatric and behavioral phenotypes and item-level GWAS for the AUDIT (13,17–26). Where possible, the largest and most recent summary statistics were used. Further information about the original studies that produced the statistics can be found in Table 1. To mitigate potential biases from population stratification, all original samples were subset to individuals of European ancestry. This study was approved by the Institutional Review Board at the University of Pittsburgh.

**Table 1:**
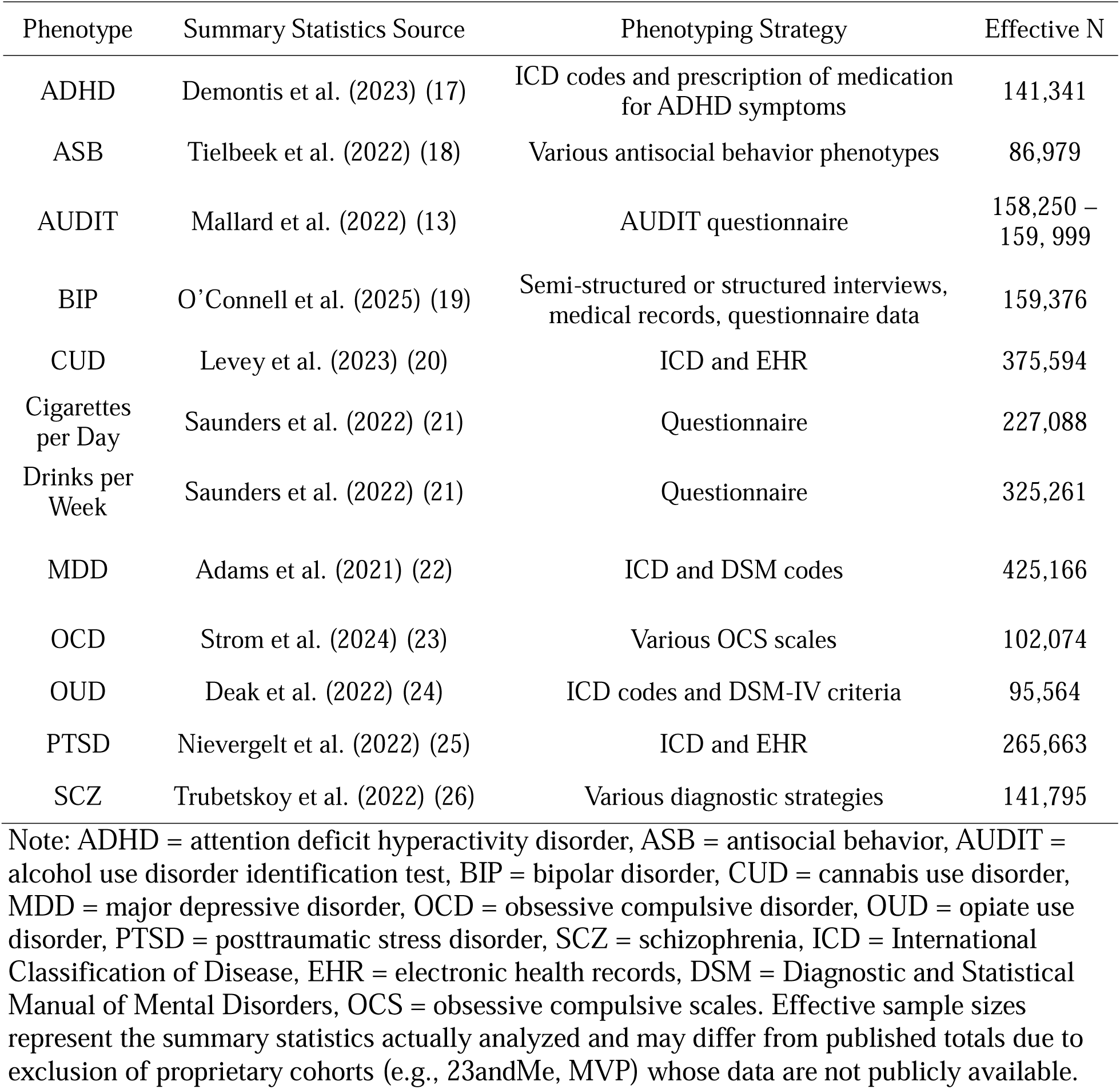
Overview of included samples.

### Data analytic plan

Analysis plans were not pre-registered. All analyses were performed using functions provided by the GenomicSEM package (14). Summary statistics were prepared for multivariate LDSC using the munge function (14), filtering to Hapmap 3 SNPs and applying default QC parameters (INFO>0.9, MAF>0.01).

#### Genetic correlation estimation

Multivariate LDSC was run using the ldsc function with the stand argument set to TRUE, producing standardized genetic covariance matrices and their associated sampling covariance matrices, along with LDSC intercepts and cross-trait intercepts. To obtain estimates of genetic correlations with the appropriate standard errors, the rgmodel function (27) was used.

#### Parallel analysis and confirmatory factor modeling (CFA)

As Qtrait analysis requires a one-factor model, we sought to test the dimensionality of AUDIT items, which were selected for inclusion based on Bonferroni-corrected significant SNP-heritability (*p*<0.05) and z-score>4. We conducted a parallel analysis on the multivariate LDSC-derived genetic correlation matrix using the paLDSC function (28). This method adapts classical parallel analysis to LDSC-derived estimates by comparing observed eigenvalues to a null distribution generated via Monte-Carlo sampling from the LDSC sampling distribution. Unlike classical principal components analysis where the number of dimensions is determined by the number of eigenvalues exceeding one, parallel analysis determines the number of important factors based on the number of eigenvalues exceeding the null threshold. We then fit a CFA based on the number of components recommended by parallel analysis using the usermodel function and all AUDIT items with statistically significant SNP-heritabilities. Model fit was assessed using standard statistical criteria: χ², CFI (≥0.90, good fit; ≥0.95, acceptable fit), and SRMR (<0.10, good fit; <0.05 acceptable fit; ((14).

#### Qtrait analysis

We performed Qtrait analysis using the Qtrait function (29) with default thresholds. The Qtrait test is a means of detecting model misfit between a “common pathway model” (CPM) and an “independent pathway model” (IPM; see Supplementary Figure 1; (14)). In the CPM, there is a single, direct path from the external correlate to the latent factor, while in the IPM, there are additional paths from the external correlate to each indicator of the latent factor. The original Qtrait test assesses the comparative fits of these models by a nested χ² difference test, where a rejection of the null hypothesis implies significant misfit of the CPM. Importantly, while the Qtrait test is a means of detecting significant deviation from the CPM, it is agnostic regarding the degree or source of the deviation. This method was recently extended to include these features (29).

Conceptually, if a given external correlate (in this study, psychiatric/behavioral phenotype) is related to the indicators of a latent variable strictly through the latent variable (i.e. as described by the CPM), then the genetic correlation between the external correlate and a given indicator would be the product of the genetic correlation between the external correlate and the latent factor and the loading of the indicator on the latent factor. Hence under the CPM, we would expect the loadings of the indicators and their correlations with an external correlate to scale approximately linearly. However, if there is indicator-specific variance that is related to the external correlate over and above what is mediated by the latent factor, the indicator(s) will deviate from CPM expectations, resulting in indicator-specific unmodeled variance in one or more of the indicators.

The means for inferring the source and degree of heterogeneity (i.e., which items result in deviations from CPM expectations), comes from an effect size measure called the local standardized root mean squared residual (lSRMR) and an outlier detection algorithm (29). Additionally, Qtrait provides an estimate of the genetic correlation between the external correlate and the latent factor after detected heterogeneity has been mitigated with direct paths between the external correlate and problematic indicators of the latent factor (i.e., outliers).

## RESULTS

### Descriptives

AUDIT items are hereafter referred to with italicized descriptive labels (see Supplementary Table 1). All AUDIT items except *Morning Drink* showed significant SNP-heritabilities and z-scores>4 (*h*^2^=0.91%-8.4%; Supplementary Table 2) and were included in genomic SEM. All external correlates had significant SNP-*h*^2^ (1.3%-19.1%; Supplementary Table 2). Results of the pairwise correlation analysis appear in Figure 1.

**Figure 1:**
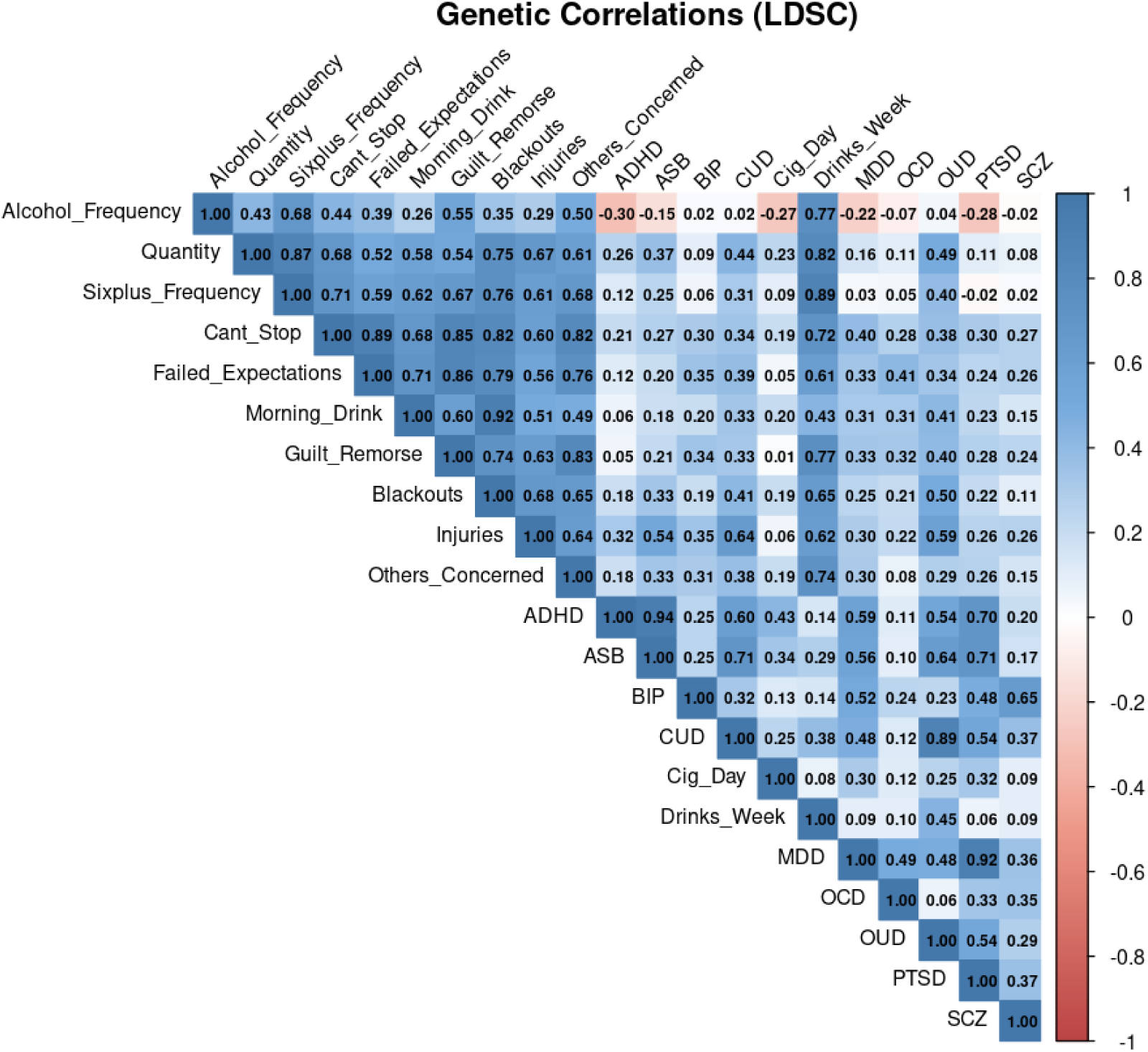
Genetic correlation matrix displaying relationships between AUDIT items and psychiatric/behavioral traits. ADHD = attention deficit hyperactivity disorder, ASB = antisocial behavior, AUDIT = alcohol use disorder identification test, BIP = bipolar disorder, CUD = cannabis use disorder, MDD = major depressive disorder, OCD = obsessive compulsive disorder, OUD = opiate use disorder, PTSD = posttraumatic stress disorder, SCZ = schizophrenia, Cig_Day= Cigarettes per day. Drinks_Week=Drinks per week. Alt text: heatmap matrix that includes all genetic correlation estimates, with each cell shaded based on statistical significance and direction of effect.

### Parallel analysis and confirmatory factor modeling

The results of the parallel analysis supported the extraction of a single factor, consistent with unidimensional structure (Supplementary Figure 2). The one-factor model composed of all AUDIT items with significant SNP-*h*^2^ fit well (χ²=322.05, *p*=2.8x10^−54^, CFI=0.95, SRMR = 0.09). All indicators loaded appreciably (range of loadings=0.58-0.92, mean loading=0.82) and significantly (all *p*’s<5.7x10^−17^) on the latent AUDIT factor (Figure 2; Supplementary Table 3).

**Figure 2:**
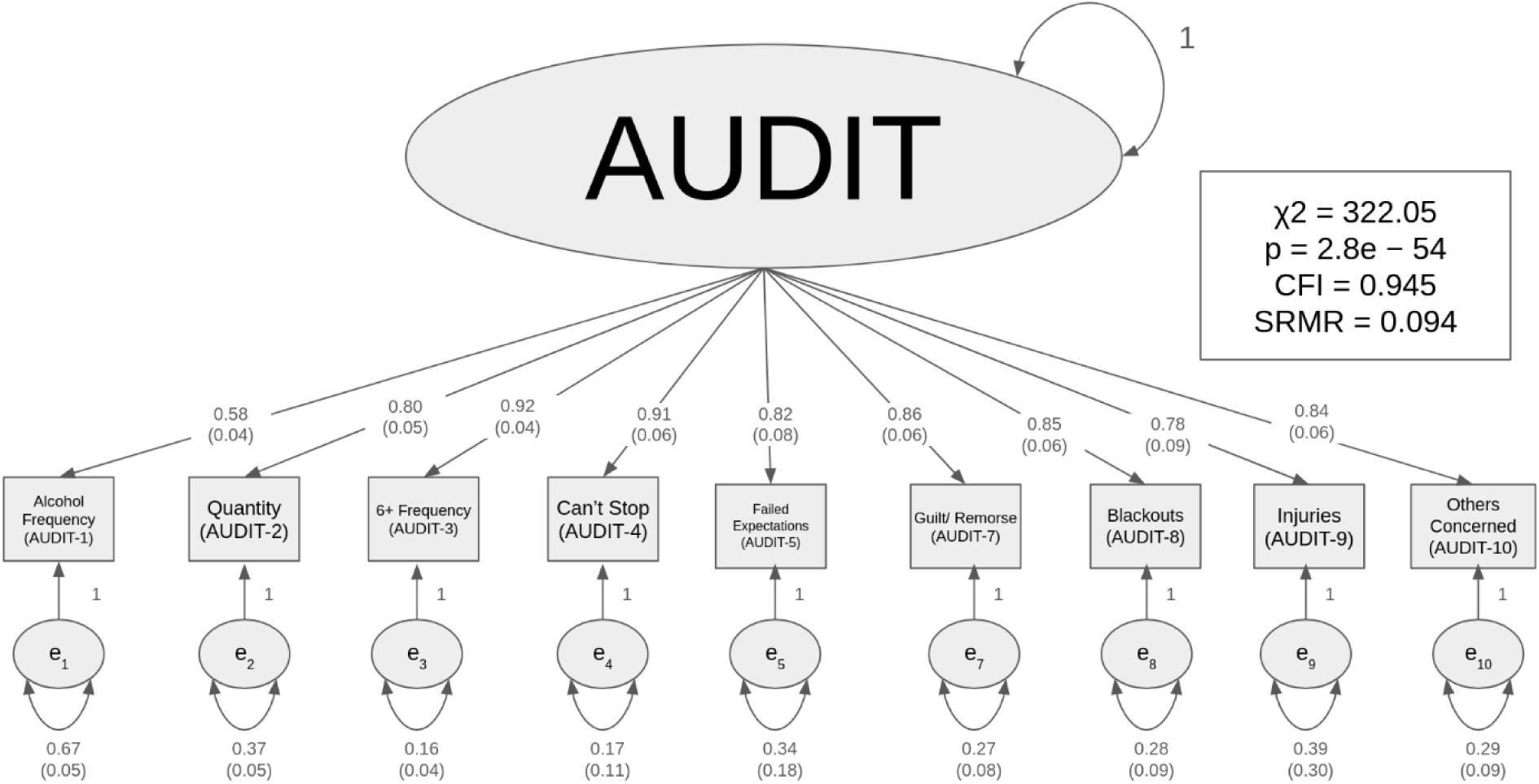
One-factor model of AUDIT items with significant SNP-heritability. Alt text: Structural equation model of one-factor model of AUDIT items that includes standardized factor loadings and model fit indices.

### Qtrait analysis

Results for the Qtrait analysis are displayed in Table 2 and Supplementary Figures 3-11. Of the 11 external correlates assessed, nine (all except cigarettes per day and OCD) displayed significant genetic correlations with the latent AUDIT factor (α_Bonferroni_=0.00455). The Qtrait statistic was significant for all nine of these external correlates, however only eight (all remaining except *Drinks per Week*) met the remaining heterogeneity criteria. Results for *Drinks per Week* (i.e., significantly genetically correlated with the PAU factor, no heterogeneity) were in line with expectations and served as basic check of the Qtrait analysis and our subsequent results.

**Table 2:**
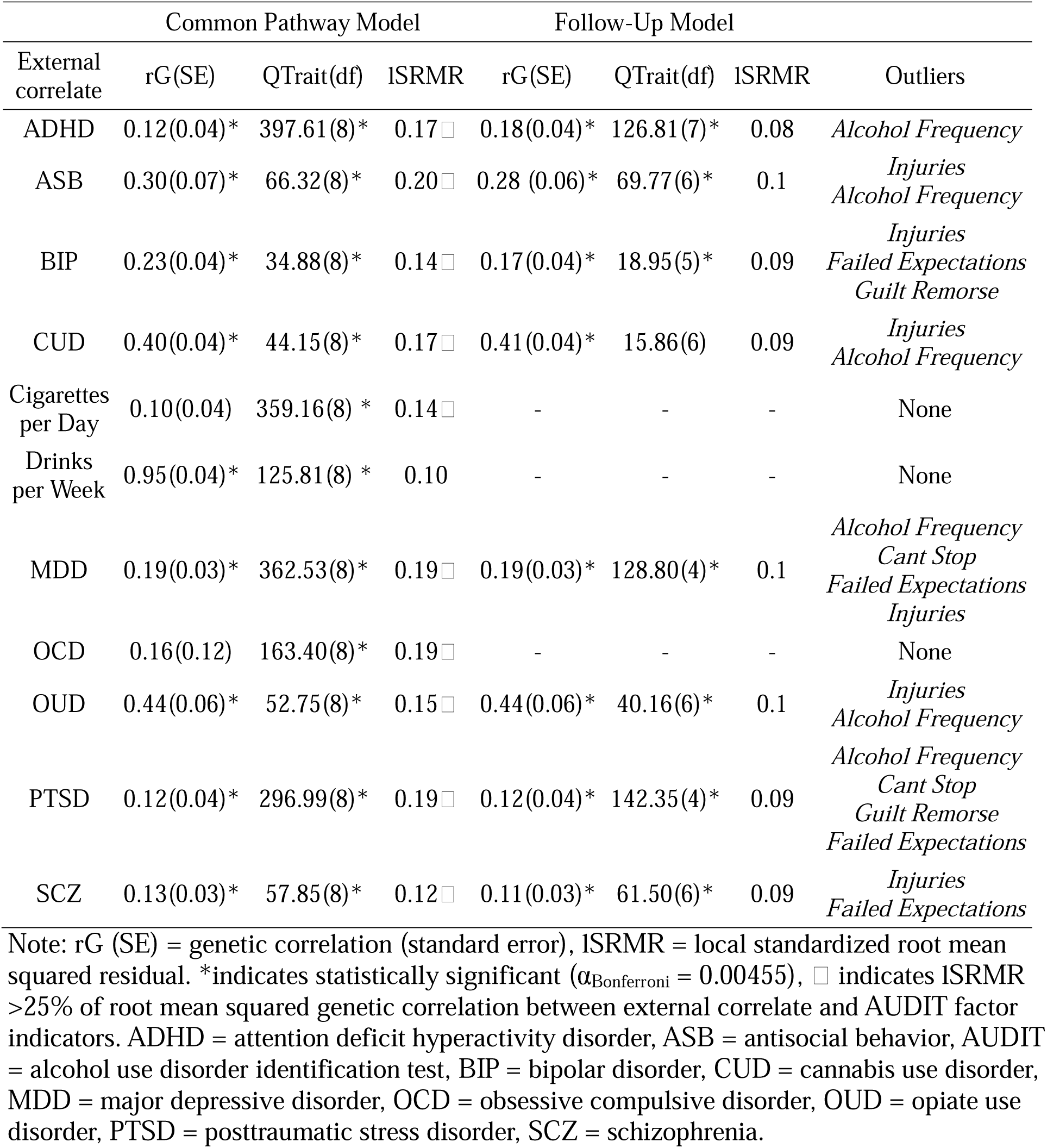
Qtrait Analysis.

The outlier detection algorithm identified specific AUDIT items that deviated from CPM expectations (Figure 3). *Alcohol Frequency* and *Injuries* were the most problematic indicators, each appearing as outliers in six of eight traits where outliers were detected. Of note, these two AUDIT items were the only indicators to display residual genetic correlation with substance use (CUD, OUD) and ASB. *Failed Expectations* was identified as an outlier in four traits (MDD, PTSD, BIP, SCZ) comprising internalizing and psychotic disorders. *Can’t Stop* and *Guilt/Remorse* were each deemed outliers with respect to two traits (MDD, PTSD) and (PTSD, BIP), respectively.

**Figure 3:**
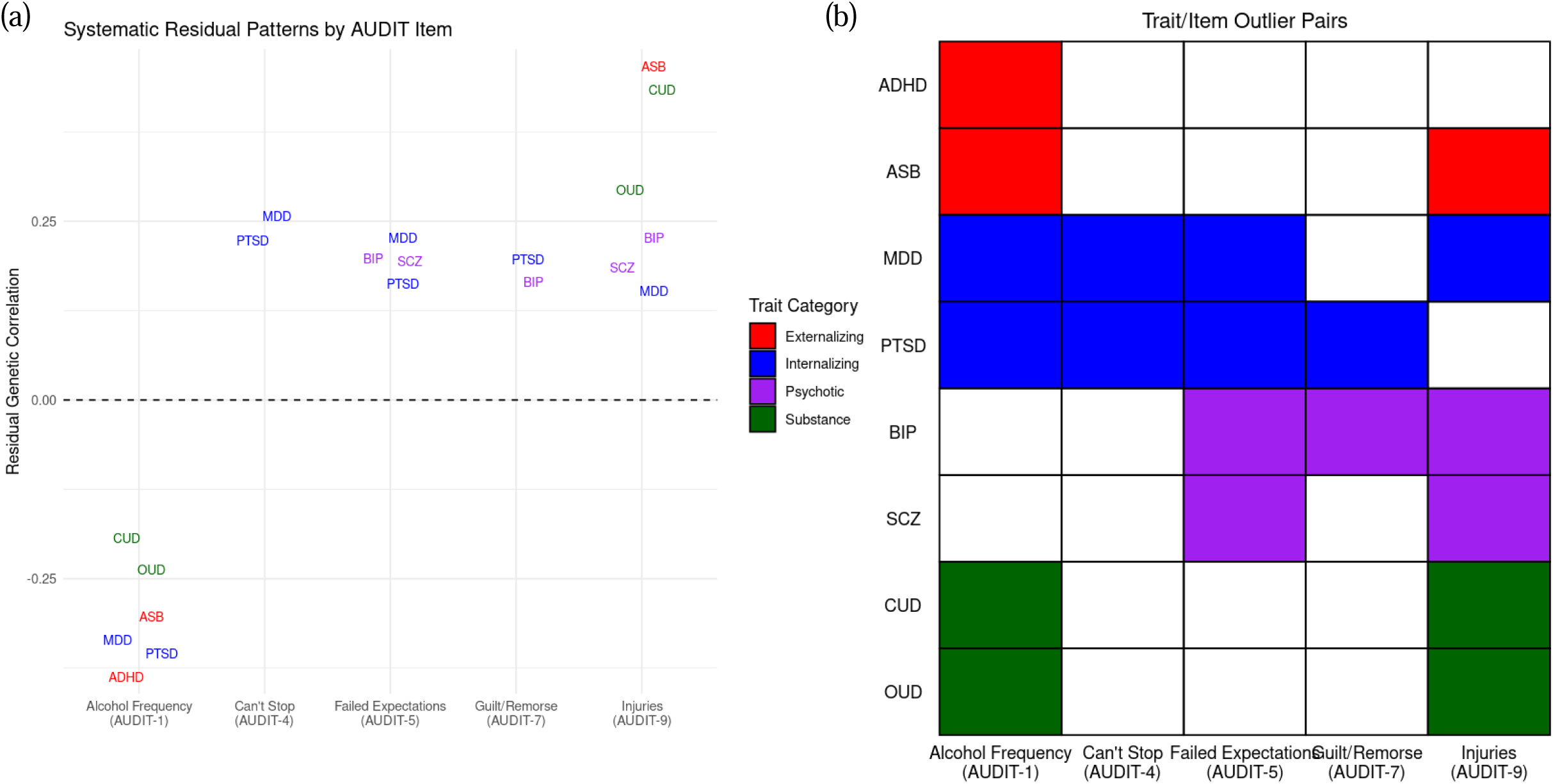
Trait/item outliers as determined by Qtrait analysis. ADHD = attention deficit hyperactivity disorder, ASB = antisocial behavior, AUDIT = alcohol use disorder identification test, BIP = bipolar disorder, CUD = cannabis use disorder, MDD = major depressive disorder, OUD = opiate use disorder, PTSD = posttraumatic stress disorder, SCZ = schizophrenia. Panel (a) shows the residual genetic correlations for each psychiatric trait/AUDIT item outlier pair. Panel (b) depicts a matrix of the varying psychiatric trait/AUDIT item outlier pairs. Alt text: Two panels to depict patterns in the residual genetic correlations between AUDIT indicators and comorbid psychiatric disorders.

With respect to the external correlates, the most problematic traits were MDD and PTSD. For both, four indicators were deemed outliers from the CPM, including *Alcohol Frequency*, *Can’t Stop*, and *Failed Expectations*. *Injuries* were detected as an additional outlier for MDD and *Guilt/Remorse* for PTSD. For BIP, three indicators were deemed outliers (*Failed Expectations*, *Guilt/Remorse*, and *Injuries*). ASB, CUD, and OUD shared an identical pair of outliers (*Alcohol Frequency* and *Injuries*), SCZ had two outliers (*Failed Expectations, Injuries)*, while the single outlier for ADHD was *Alcohol Frequency*.

The genetic correlations between external traits and the AUDIT factor in the CPM and FUM showed varying patterns of change: some increased (e.g., ADHD: 0.12 to 0.18), others decreased (e.g., BIP: 0.23 to 0.17), and some remained largely unchanged (e.g., CUD: 0.40 to 0.41; PTSD: 0.12 to 0.12). These differences were assessed using Fisher’s Z-transformation test. None were statistically significant (all *p*’s >0.25).

Examination of the residual genetic correlations for identified outlier indicators revealed systematic directional patterns across external correlates (Figure 3). *Alcohol Frequency* consistently exhibited negative residuals when identified as an outlier (ranging from -0.37 to - 0.21), indicating weaker-than-expected genetic associations with external correlates relative to the CPM. Conversely, all other indicators consistently showed positive residuals (ranging from 0.15 to 0.45), suggesting stronger-than-expected associations relative to the CPM. The three largest residual correlations were all with respect to *Injuries* (ASB=0.45, CUD=0.42, OUD=0.32), while the seven largest (in terms of absolute value) were comprised of these three values and four others which were all with respect to *Alcohol Frequency* (ADHD=-0.37, PTSD=- 0.35, MDD=-0.33, ASB=-0.32).

## DISCUSSION

This study examined the extent to which individual symptoms of PAU contributed to the observed genetic comorbidity between PAU and other psychiatric disorders. We found that the majority of PAU indicators, particularly those relating to *consequences* from alcohol use, showed unique associations with various comorbid psychiatric conditions across disorder spectra over and above the common PAU factor. Moreover, a large majority of comorbid psychiatric conditions (nine of eleven) were implicated in these associations. Modeling these associations shifted some genetic correlations between psychiatric conditions and PAU. Results suggest that certain AUDIT items capture genetic variance that is not fully represented by the general AUDIT liability factor, potentially reflecting distinct biological pathways or measurement characteristics.

We found that PAU indicators reflecting negative psychosocial and health consequences of alcohol use, specifically *Injuries*, *Failed expectations*, and *Guilt/Remorse* showed positive residual genetic associations with six, four, and two comorbid psychiatric conditions, respectively, and these effects did not operate through the common PAU factor. The effects on *Injuries*, *Failed expectations,* and *Guilt/Remorse* were each distributed across multiple types of comorbid disorders (e.g., internalizing *and* psychotic disorders). From this we surmise that findings reflect conceptual and measurement limitations associated with using alcohol-related consequences to operationalize PAU rather than biologically meaningful shared genetic pathways.

Indeed, because alcohol consequences are multiply determined and not easily attributable to alcohol use alone, researchers have argued that using them to assess AUD could artifactually inflate rates of psychiatric comorbidity (9,11). For example, many psychiatric conditions, such as MDD and BIP, are diagnosed only if individuals exhibit impairments in social, occupational or other important areas of functioning due to symptoms. For individuals with functional impairments, infractions related to alcohol use might have more serious repercussions, perhaps leading to endorsement of alcohol-related consequences even in the relative absence of other alcohol-related pathology. Third variables could also explain these associations. As one example, avoidant coping styles have been shown to increase risk for the onset and maintenance of MDD and PTSD while also increasing risk for drinking to avoid stressful responsibilities (30–32). This third variable could cause alcohol-related psychosocial and health consequences even when few other signs of PAU are evident.

Notably, *Injuries* showed the most numerous specific direct associations with comorbid psychiatric conditions. It was also the only indicator with more positive-than-expected associations with substance use disorders and ASB, and these showed the largest residual genetic correlations observed in this study. Thus, although hazardous alcohol use in part reflects genetic variation common to PAU, it also captures genetic variation that is unique. Perhaps this unique genetic variation influences trait-level impulsivity or rashness that influences other psychiatric conditions as well as hazardous alcohol use independently from other PAU symptoms. Items assessing hazardous use may thus be especially likely to artifactually inflate genetic correlation between PAU and other psychiatric disorders. While our results replicate prior research (10–12), our study—to the best of our knowledge—is the first to provide evidence that shared genetic liability between alcohol-related consequences and comorbid psychiatric conditions contributes to their phenotypic covariance. These findings also add to interpretations that trait-level genetic factors contribute to these associations.

The PAU indicator, *Can’t Stop*, is intended to capture impaired control over drinking (6) and was related to MDD and PTSD. Because *Can’t Stop* showed associations with only internalizing disorders that are known to be tightly genetically linked (2), it is possible that a substantive genetic pathway explains these associations. Speculatively, perhaps deficiencies in the experience of reward that are common to MDD and PTSD represent a unique genetic pathway to impaired control over alcohol use independent of genetic variation that is common to PAU.

The indicator, *Alcohol Frequency*, showed *negative* residual genetic associations with CUD, OUD, ADHD, ASB, MDD and PTSD. These findings are consistent with a larger literature showing that associations between the frequency of alcohol use and psychiatric diagnoses (e.g., PTSD, MDD, ADHD) were near zero or small and negative, but became significantly negative when partialling out the effects of alcohol problems (33–35). Thus, after controlling for genetic variance shared with problematic alcohol use, drinking alcohol frequently may reflect having more leisure time to enjoy alcohol, having the resources to obtain alcohol, or using alcohol to celebrate happy occasions or to socialize (qualities likely to be inversely associated with psychiatric disorders). Indeed, life satisfaction was shown to share genetic etiology with typical alcohol use once problem use was partialled out (36).

When comparing results from the common pathway to the follow up models, we found that the genetic correlations between the common PAU factor and psychiatric conditions generally showed small changes (all changes in *r*<0.06) and none were statistically significant. At first glance, this might appear to suggest that including alcohol-related consequences in measures of PAU may not substantially affect estimates of genetic correlation. Consider, however, that many genetically informative studies use AUD *diagnoses* as an outcome, for which only two symptoms need to be met. Depending on the extent to which endorsements of alcohol-related consequences increase the odds of AUD diagnosis, estimates of genetic correlation between other psychiatric disorders and AUD could be substantially impacted by the inclusion of alcohol-related consequences. Similarly, many genetically informative studies use as an outcome AUD or AUDIT symptom counts in which each indicator is weighted approximately equally. This could result in more biased genetic correlations than observed here, in which PAU was measured using empirically derived weights via factor analysis. Moreover, if alcohol-related consequences reflect something *besides* alcohol-related pathology (i.e., functional impairment from other psychiatric disorders), including them in measures of PAU and AUD could introduce noise into studies on the etiology of AUD alone. It will be important to continue to explore these questions in future studies.

If alcohol-related consequences introduce some confounding into PAU measures, which symptoms might be less biased representations of PAU? We found that *Alcohol Quantity*, *6+ Frequency*, *Blackouts*, and *Others Concerned* did not show specific direct associations with any other psychiatric conditions. In contrast to the alcohol consequence items, strong endorsements of *Alcohol Quantity*, *6+ Frequency*, *Blackouts* are predicated on the consumption of large quantities of alcohol and *Others concerned* might tap a higher severity of symptomatology. PAU symptoms that are more directly related to the consumption of alcohol may be less likely to exhibit the same measurement limitations associated with alcohol-related consequences.

This study had limitations. Analyses relied on summary statistics derived from population GWAS and use LDSC to estimate SNP-heritabilities and genetic correlations. These are known to have potential biasing factors, including within- and cross-trait assortative mating, population stratification, indirect genetic effects and ascertainment/participation bias, and LD-dependent architecture of the traits in question (37–40). Unfortunately, we did not have access to within-family GWAS data on our traits of interest, which would have mitigated some of these biases. We attempted to mitigate effects of population stratification by using LDSC and restricting our sample to individuals of European ancestry. However, this strategy limits the generalizability of our findings and thus should be replicated with more diverse samples and robust methods when adequate sample sizes become available. Additionally, as many of our summary statistics came from meta-analyses of cohorts with various phenotyping strategies, some degree of measurement heterogeneity is expected. Furthermore, the AUDIT, though widely used, is not directly parallel with AUD criteria, necessitating studies on AUD diagnosis in the future.

In conclusion, using genomic data we provide evidence to support the notion that alcohol-related consequences, including *Injuries, Failed expectations,* and *Guilt/Remorse*, may unduly reflect dysfunction from comorbid psychiatric conditions or related third variables. We find that *Alcohol Quantity*, *6+ Frequency*, *Blackouts*, and *Others concerned* might be less confounded representations of PAU. Findings demonstrate the need for future work to study the extent to which including alcohol-related consequences in diagnoses of AUD could affect studies on psychiatric comorbidity and the etiology of AUD itself.

## Data Availability

All data produced are available online at the psychiatric genetics consortium, CTG Lab website, University of Minnestoa site, and the Gelertner Lab site.

https://medicine.yale.edu/lab/gelernter/stats/

https://conservancy.umn.edu/items/91f6a003-6af2-4809-9785-53dc579dc788

https://cncr.nl/research/summary_statistics/

https://pgc.unc.edu/for-researchers/download-results/

