## Supplementary Materials for "Symptoms of problematic alcohol use differ in their genetic associations with comorbid internalizing, externalizing, and neurodevelopmental psychiatric disorders"

Supplementary Table 1: AUDIT Questionnaire items and shorthand labels used in text.

| AUDIT Item | Item Label | Full Question Text |
| --- | --- | --- |
| AUDIT-1 | Alcohol Frequency | How often do you have a drink containing alcohol? |
| AUDIT-2 | Quantity | How many drinks containing alcohol do you have on a typical day when you are drinking? |
| AUDIT-3 | 6+ Frequency | How often do you have six or more drinks on one occasion? |
| AUDIT-4 | Can't Stop | How often during the last year have you found that you were not able to stop drinking once you had started? |
| AUDIT-5 | Failed Expectations | How often during the last year have you failed to do what was normally expected from you because of drinking? |
| AUDIT-6 | Morning Drink | How often during the last year have you needed a first drink in the morning to get yourself going after a heavy drinking session? |
| AUDIT-7 | Guilt/Remorse | How often during the last year have you had a feeling of guilt or remorse after drinking? |
| AUDIT-8 | Blackouts | How often during the last year have you been unable to remember what happened the night before because you had been drinking? |
| AUDIT-9 | Injuries | Have you or someone else been injured as a result of your drinking? |
| AUDIT-10 | Others Concerned | Has a relative or friend or a doctor or another health worker been concerned about your drinking or suggested you cut down? |

Supplementary Table 2: SNP-heritability estimates, effective sample sizes and intercepts of all traits analyzed.

| Info | Trait | *h*^2^ SNP (SE) | *h*^2^ SNP z-score | *h*^2^ SNP *p* | Neff | Intercept |
| --- | --- | --- | --- | --- | --- | --- |
| Alcohol Frequency | AUDIT 1 | 0.0843 (0.0049) | 17.13 | 1.97E-64 | 159199 | 1.0082 |
| Quantity | AUDIT 2 | 0.0494 (0.0032) | 14.86 | 1.29E-48 | 158250 | 1.0045 |
| 6+ Frequency | AUDIT 3 | 0.0595 (0.0033) | 26.85 | 1.78E-157 | 158779 | 1.0063 |
| Can't Stop | AUDIT 4 | 0.0176 (0.0029) | 6.04 | 3.34E-08 | 158801 | 1.0076 |
| Failed Expectations | AUDIT 5 | 0.0114 (0.0027) | 4.26 | 4.28E-04 | 158785 | 1.0039 |
| Morning Drink | AUDIT 6 | 0.0035 (0.0022) | 1 | 1.00E+00 | 158803 | 0.9947 |
| Guilt/Remorse | AUDIT 7 | 0.0252 (0.0030) | 8.9 | 1.20E-17 | 159030 | 1.0124 |
| Blackouts | AUDIT 8 | 0.0266 (0.0028) | 11.19 | 9.21E-28 | 158797 | 0.9991 |
| Injuries | AUDIT 9 | 0.0091 (0.0026) | 4.04 | 1.12E-03 | 159062 | 1.0065 |
| Others Concerned | AUDIT 10 | 0.0272 (0.0029) | 10.89 | 2.84E-26 | 159076 | 1.0065 |
| Externalizing/  Neurodevelopmental | ADHD | 0.1449 (0.0037) | 66.34 | 0.00E+00 | 141341 | 1.0253 |
| Externalizing/  Neurodevelopmental | ASB | 0.0748 (0.0049) | 21.48 | 5.15E-101 | 86979 | 1.0159 |
| Psychotic | BIP | 0.1181 (0.0035) | 75.65 | 0.00E+00 | 159376 | 1.0551 |
| Substance | CUD | 0.0554 (0.0024) | 27.61 | 1.78E-166 | 375594 | 0.993 |
| Substance | Cigarettes per Day | 0.0742 (0.0027) | 38.22 | 0.00E+00 | 227088 | 1.0655 |
| Substance | Drinks per Week | 0.0546 (0.0028) | 29.48 | 9.79E-190 | 325261 | 1.0931 |
| Internalizing | MDD | 0.0617 (0.0020) | 20.71 | 5.38E-94 | 425166 | 1.0597 |
| Internalizing | OCD | 0.0134 (0.0035) | 5.7 | 2.46E-07 | 102074 | 1.0044 |
| Substance | OUD | 0.1007 (0.0050) | 48.36 | 0.00E+00 | 95564 | 1.0102 |
| Internalizing | PTSD | 0.0548 (0.0023) | 18.34 | 8.44E-74 | 265663 | 1.0296 |
| Psychotic | SCZ | 0.1909 (0.0035) | 27.57 | 4.89E-166 | 141795 | 1.078 |

Note. *p*-value is Bonferroni-corrected.

| Indicator | β | CI_95_ |
| --- | --- | --- |
| AUDIT_1 | 0.58 | (0.5,0.66) |
| AUDIT_2 | 0.8 | (0.7,0.89) |
| AUDIT_3 | 0.92 | (0.84,0.99) |
| AUDIT_4 | 0.91 | (0.78,1.04) |
| AUDIT_5 | 0.82 | (0.65,0.98) |
| AUDIT_7 | 0.86 | (0.74,0.97) |
| AUDIT_8 | 0.85 | (0.74,0.96) |
| AUDIT_9 | 0.78 | (0.6,0.97) |
| AUDIT_10 | 0.84 | (0.73,0.96) |

Supplementary Table 3. AUDIT One-Factor Model Confidence Intervals

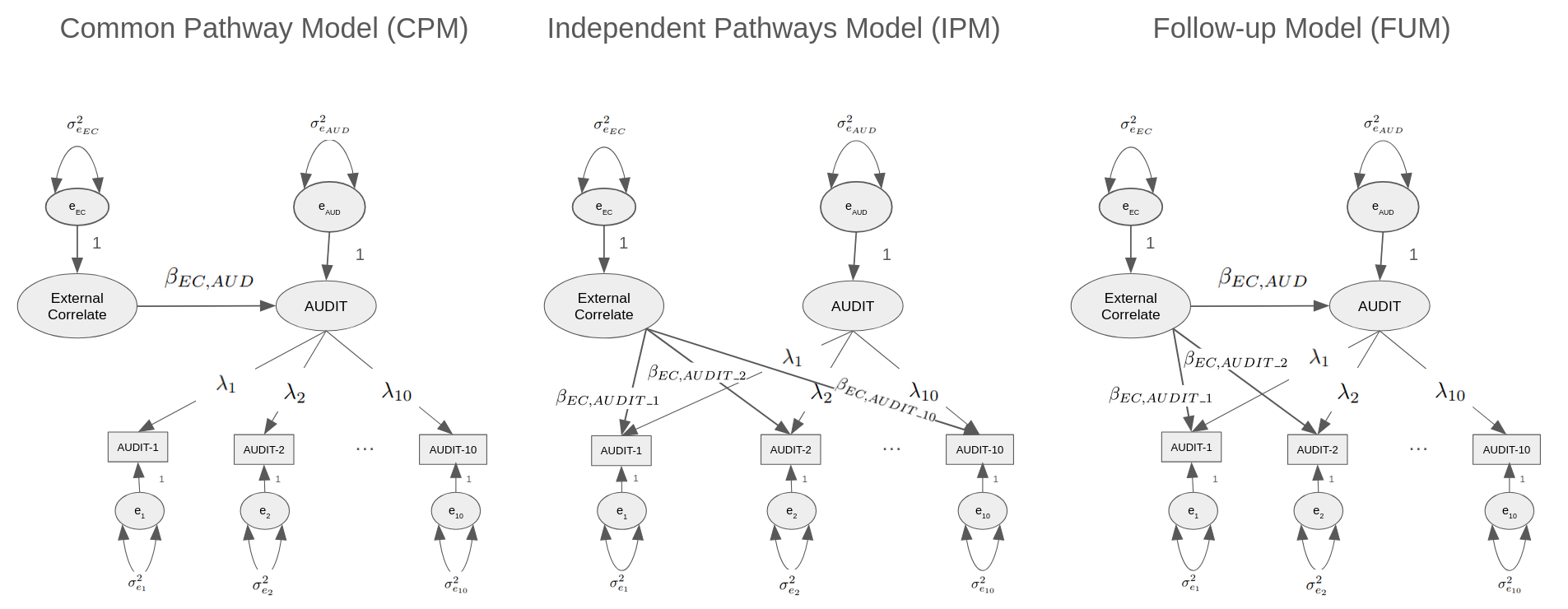

Supplementary Figure 1: Conceptual schematic of common pathway model (CPM), independent pathways model (IPM), and follow-up model (FUM)

(a)
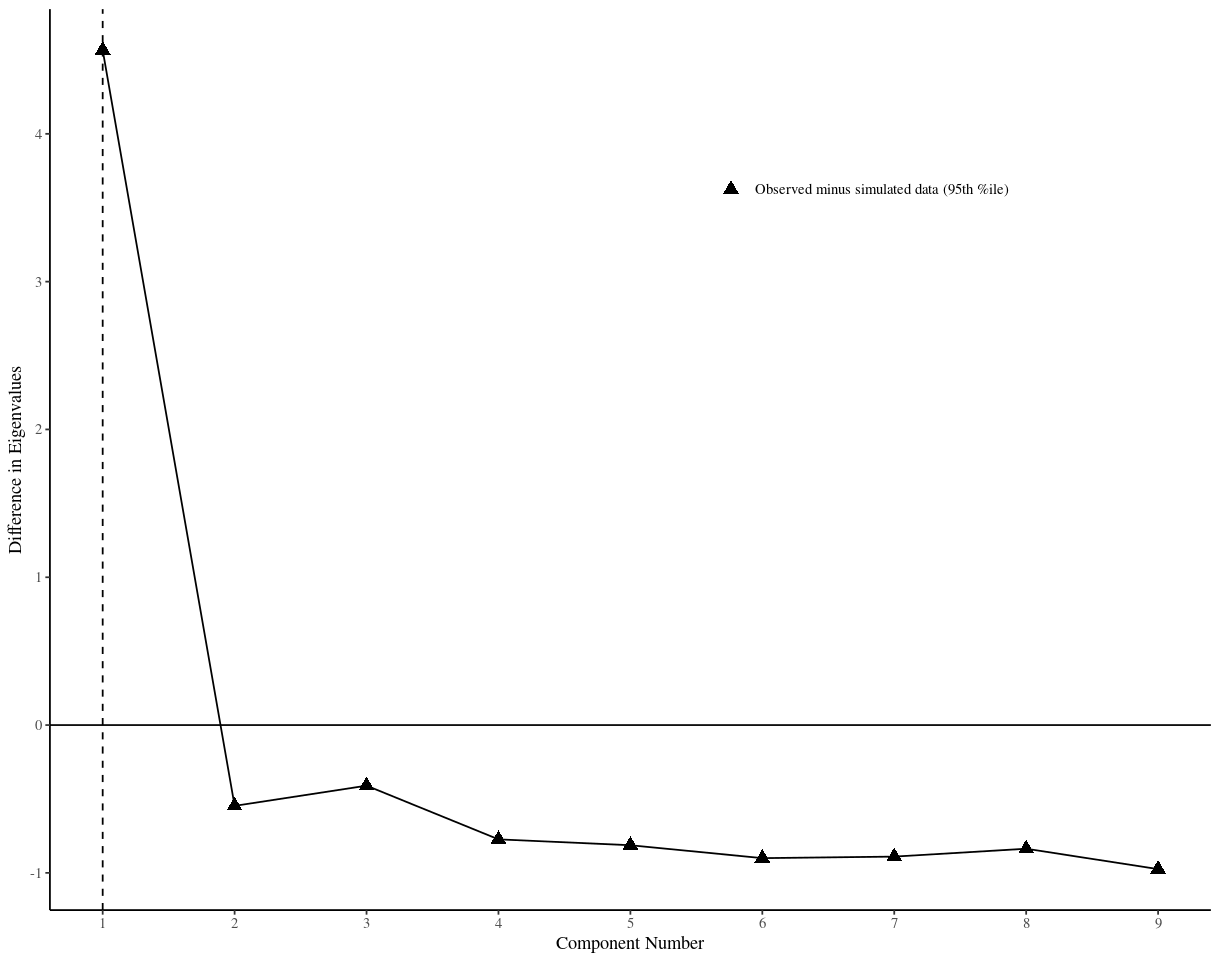

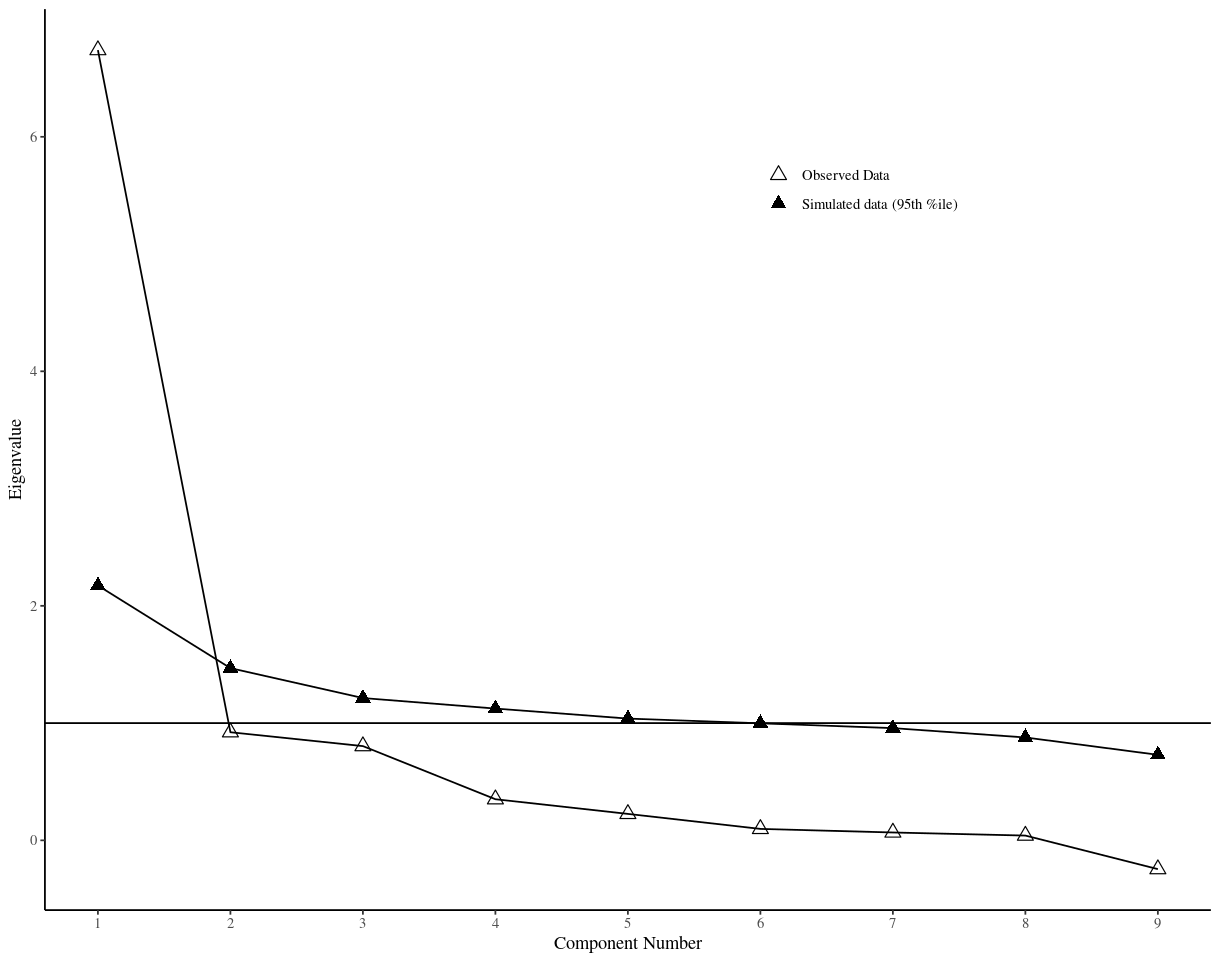
 (b)

Supplementary Figure 2: Parallel Analysis of AUDIT items with significant SNP-heritability (observed vs simulated). Panel (a) shows observed vs simulated data. Panel (b) shows observed minus simulated data.

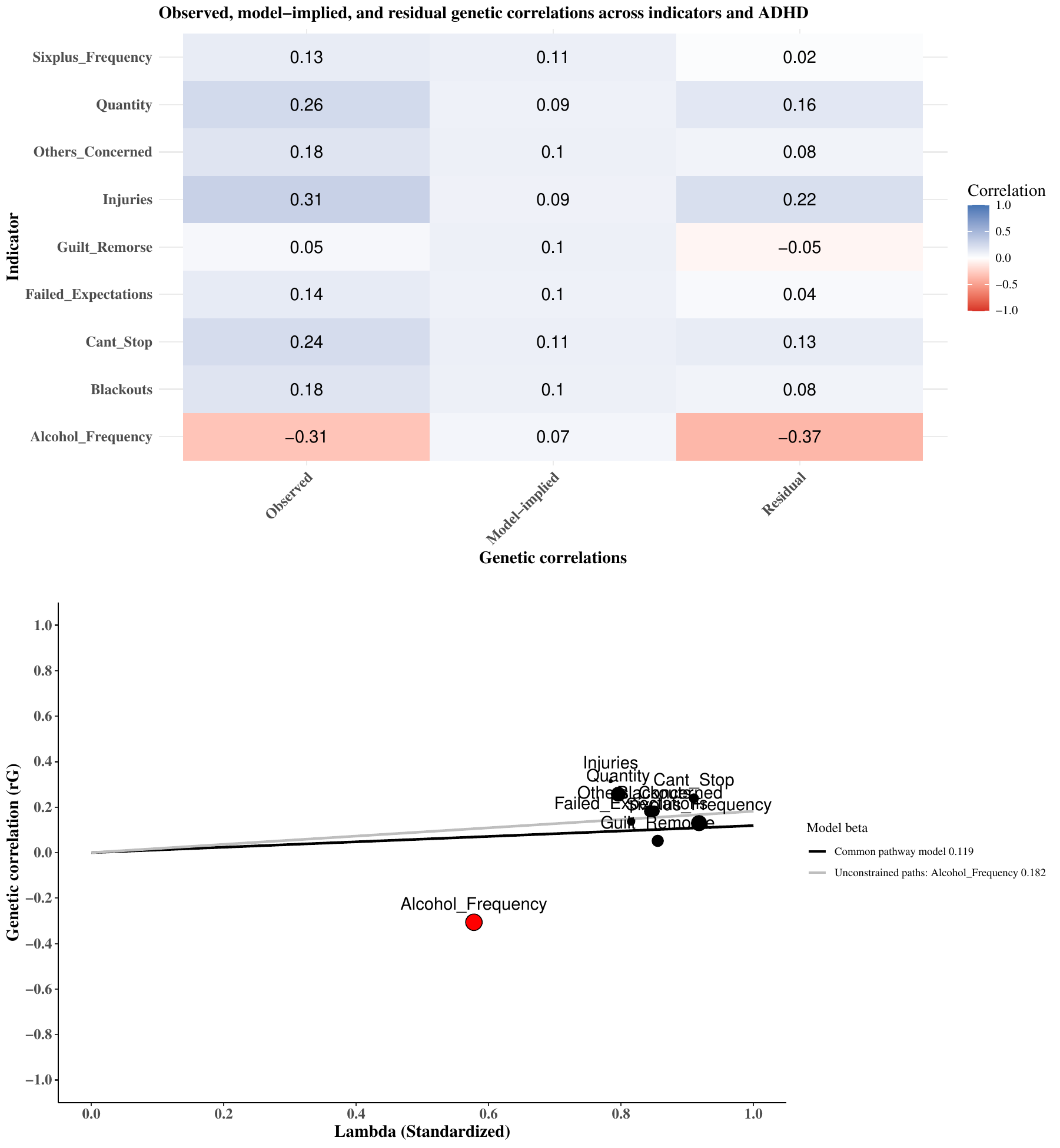

Supplementary Figure 3: Qtrait analysis of ADHD

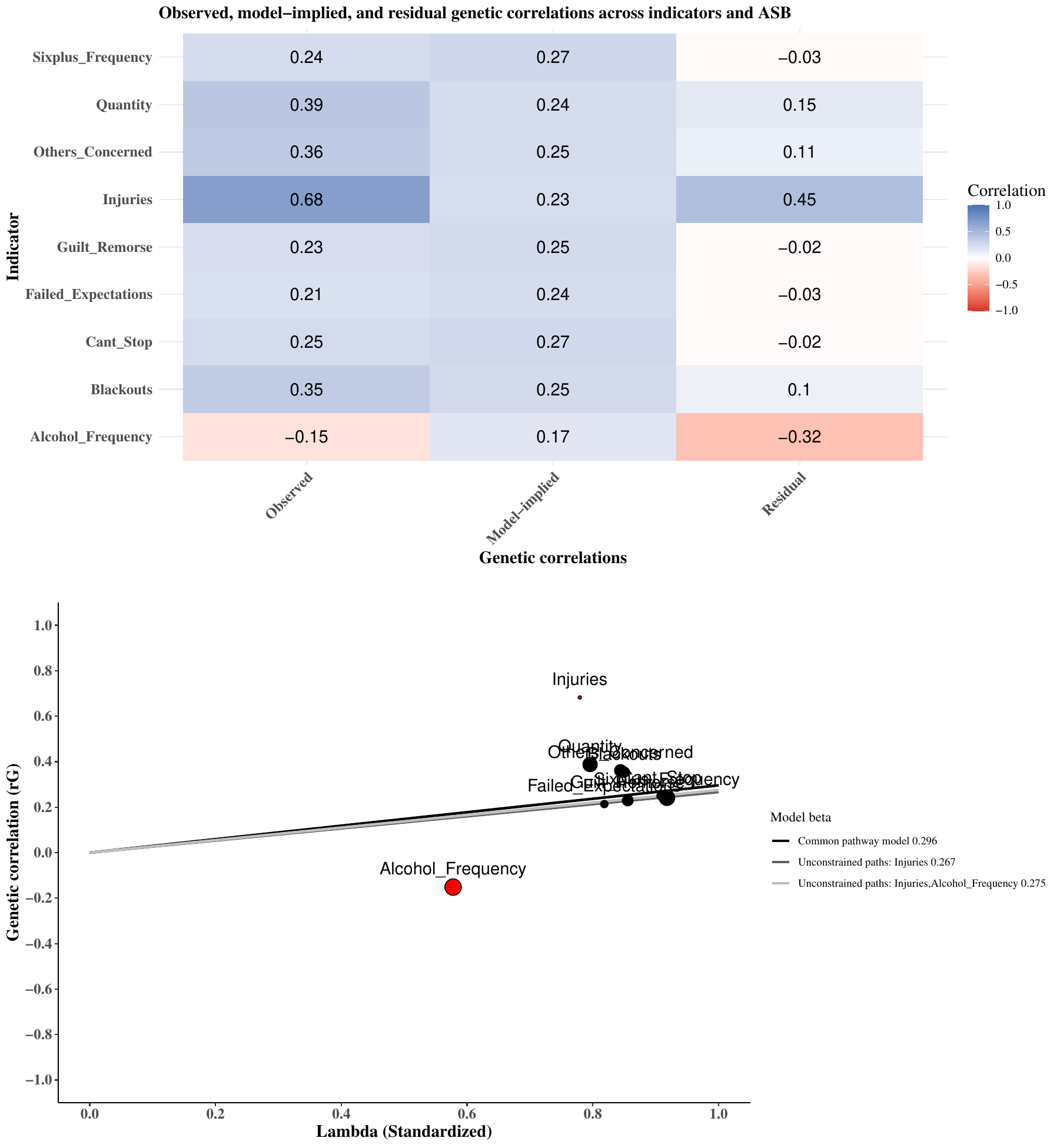

Supplementary Figure 4: Qtrait analysis of ASB

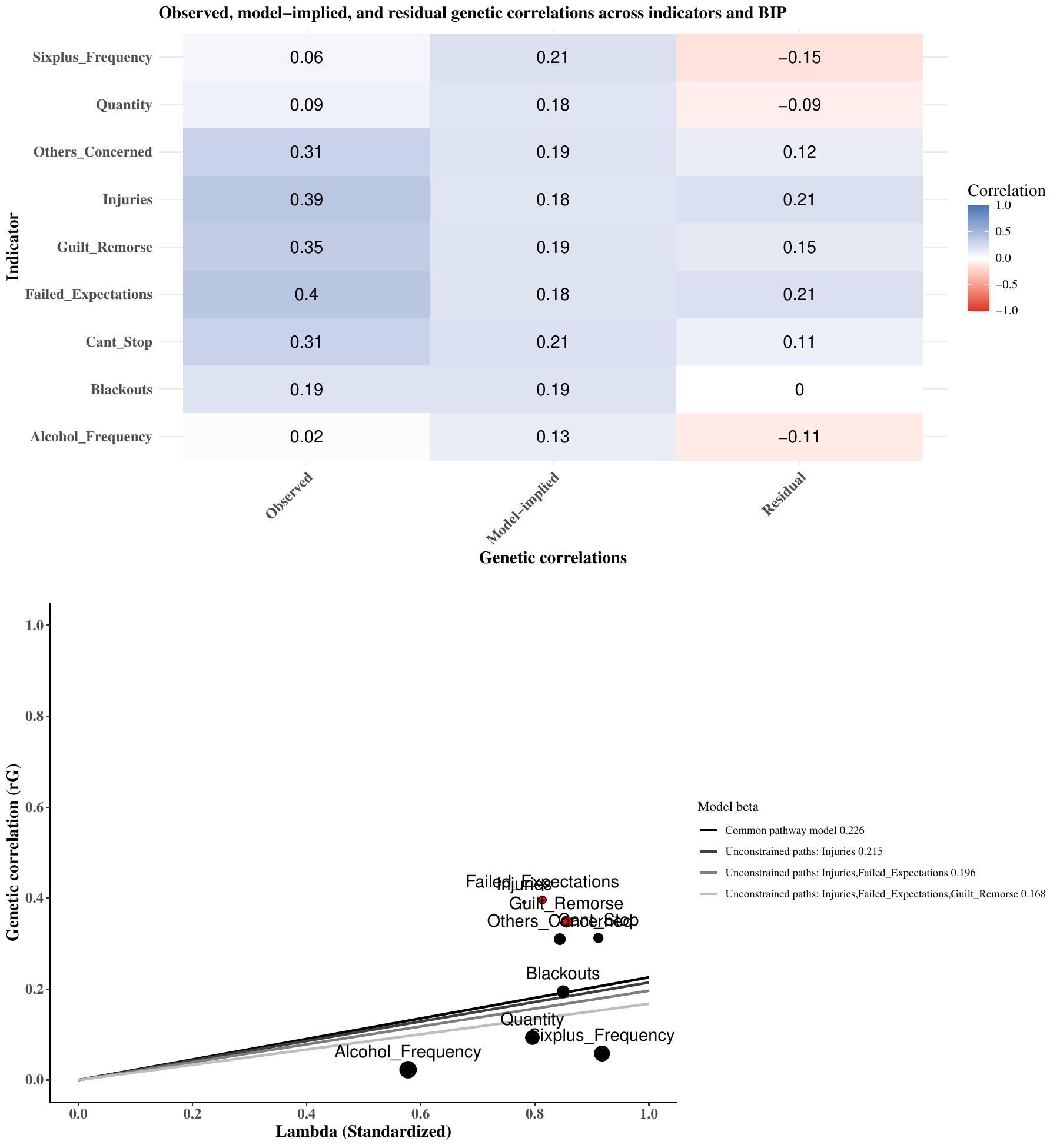

Supplementary Figure 5: Qtrait analysis of BIP

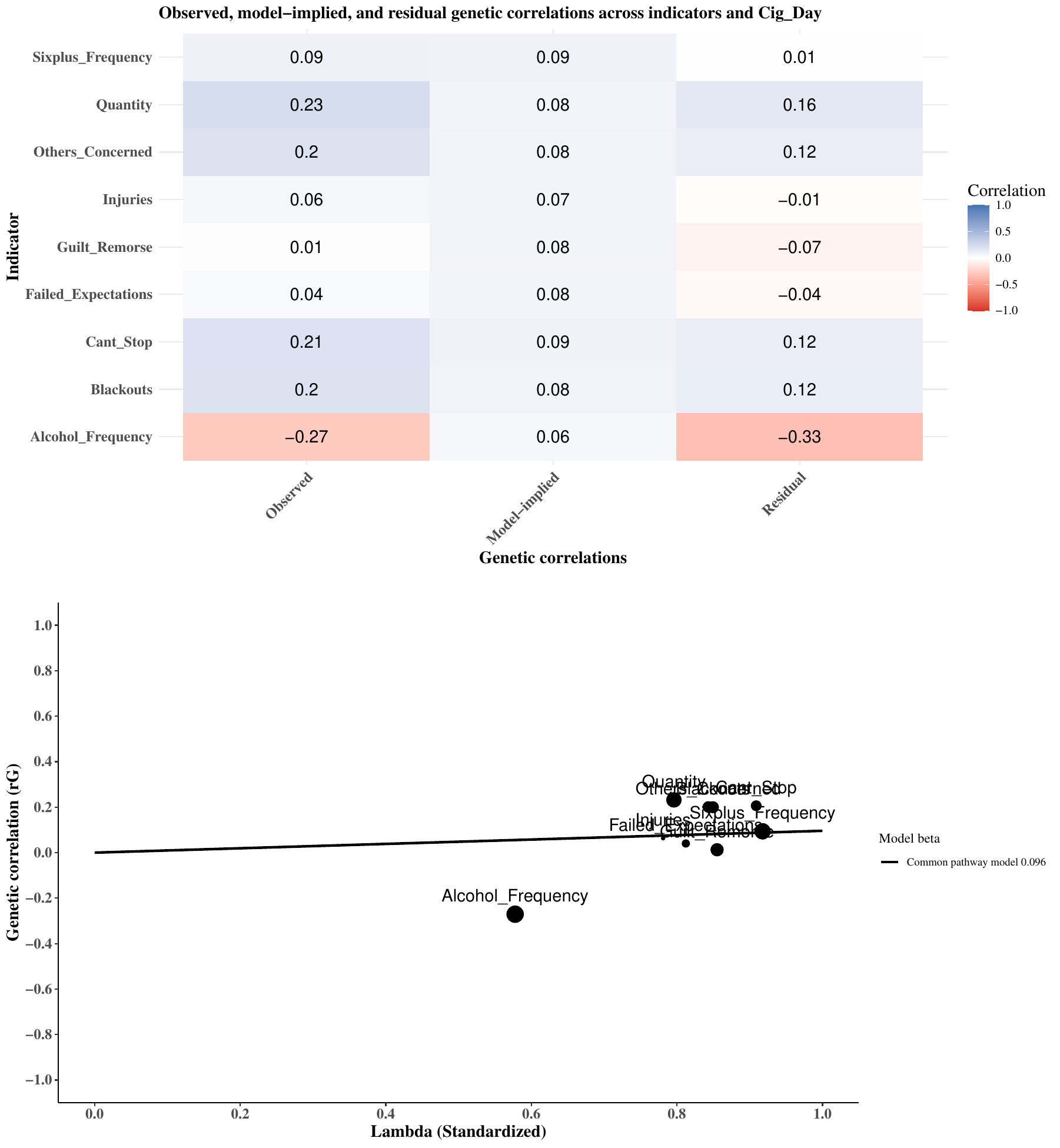

Supplementary Figure 6: Qtrait analysis of Cigarettes per Day

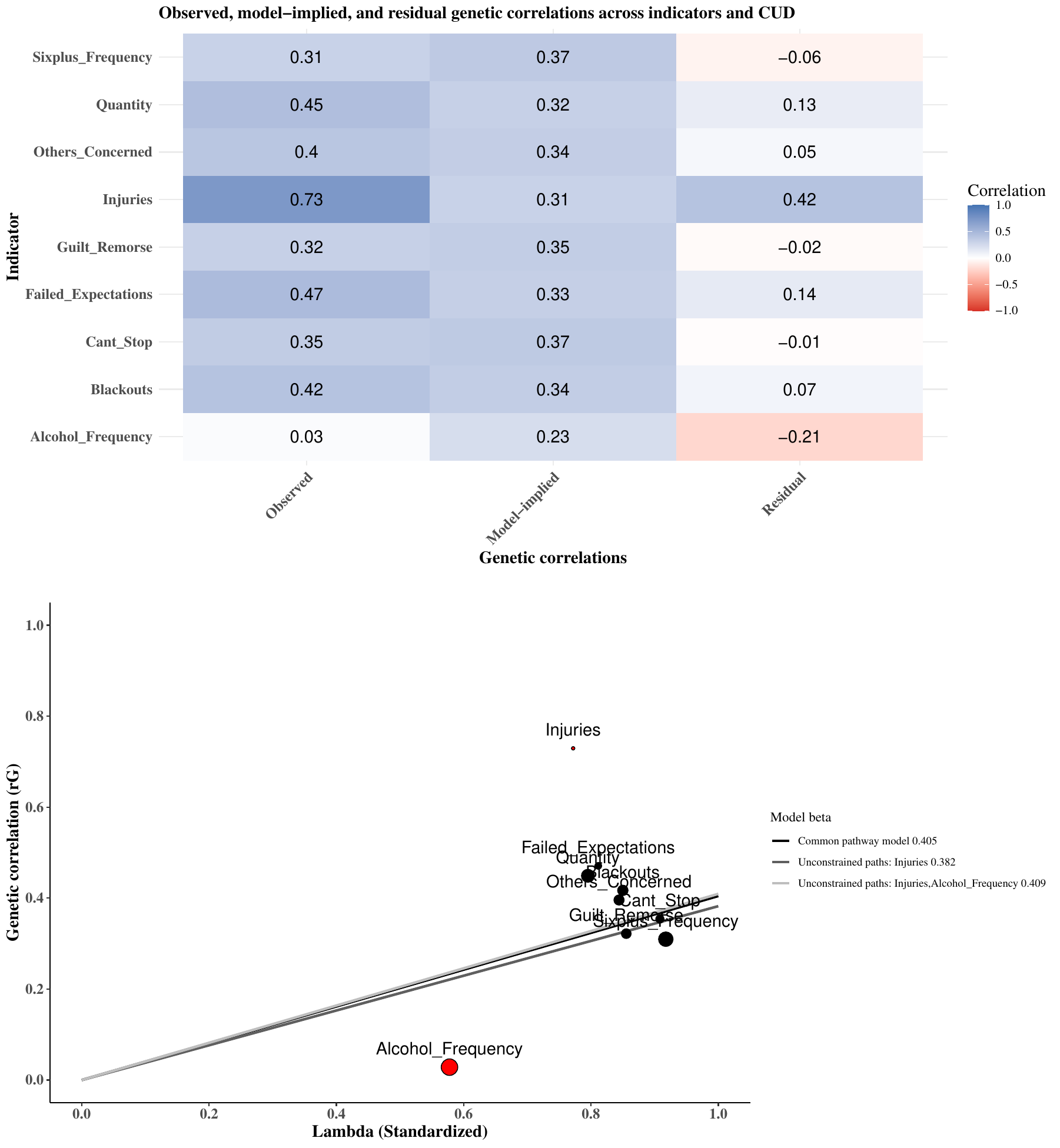

Supplementary Figure 7: Qtrait analysis of CUD

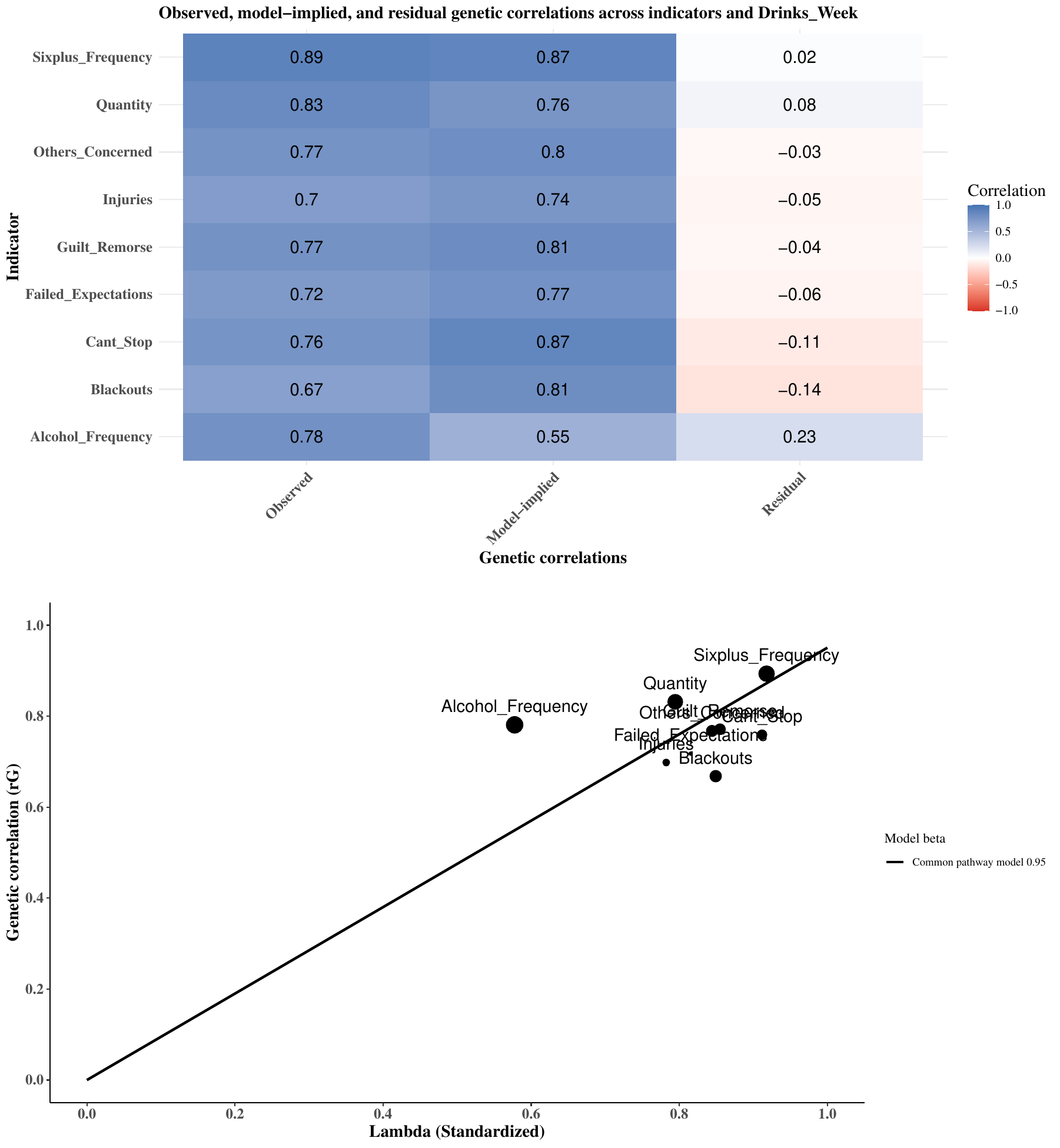

Supplementary Figure 8: Qtrait analysis of Drinks per Week

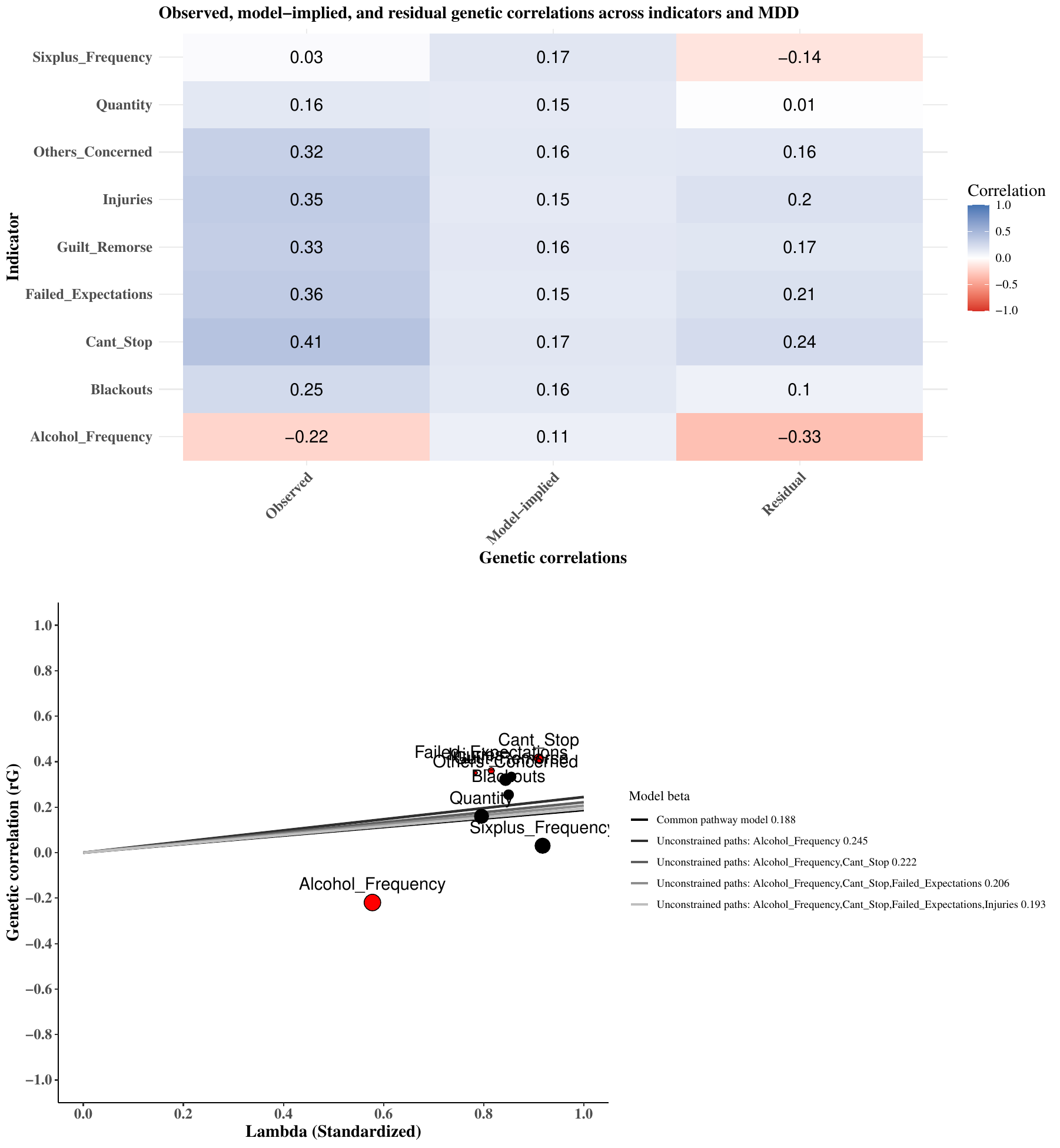

Supplementary Figure 9: Qtrait analysis of MDD

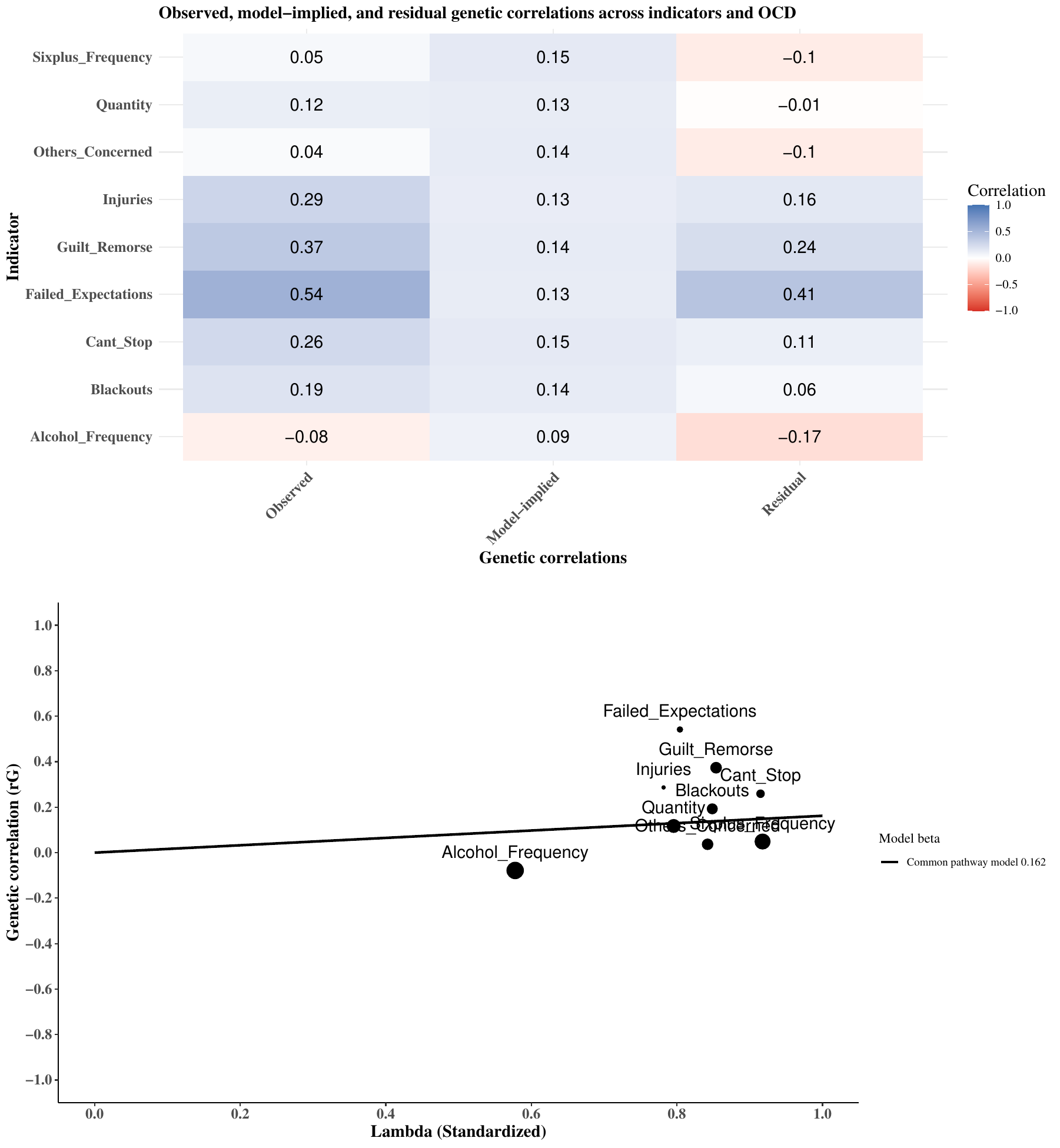

Supplementary Figure 10: Qtrait analysis of OCD

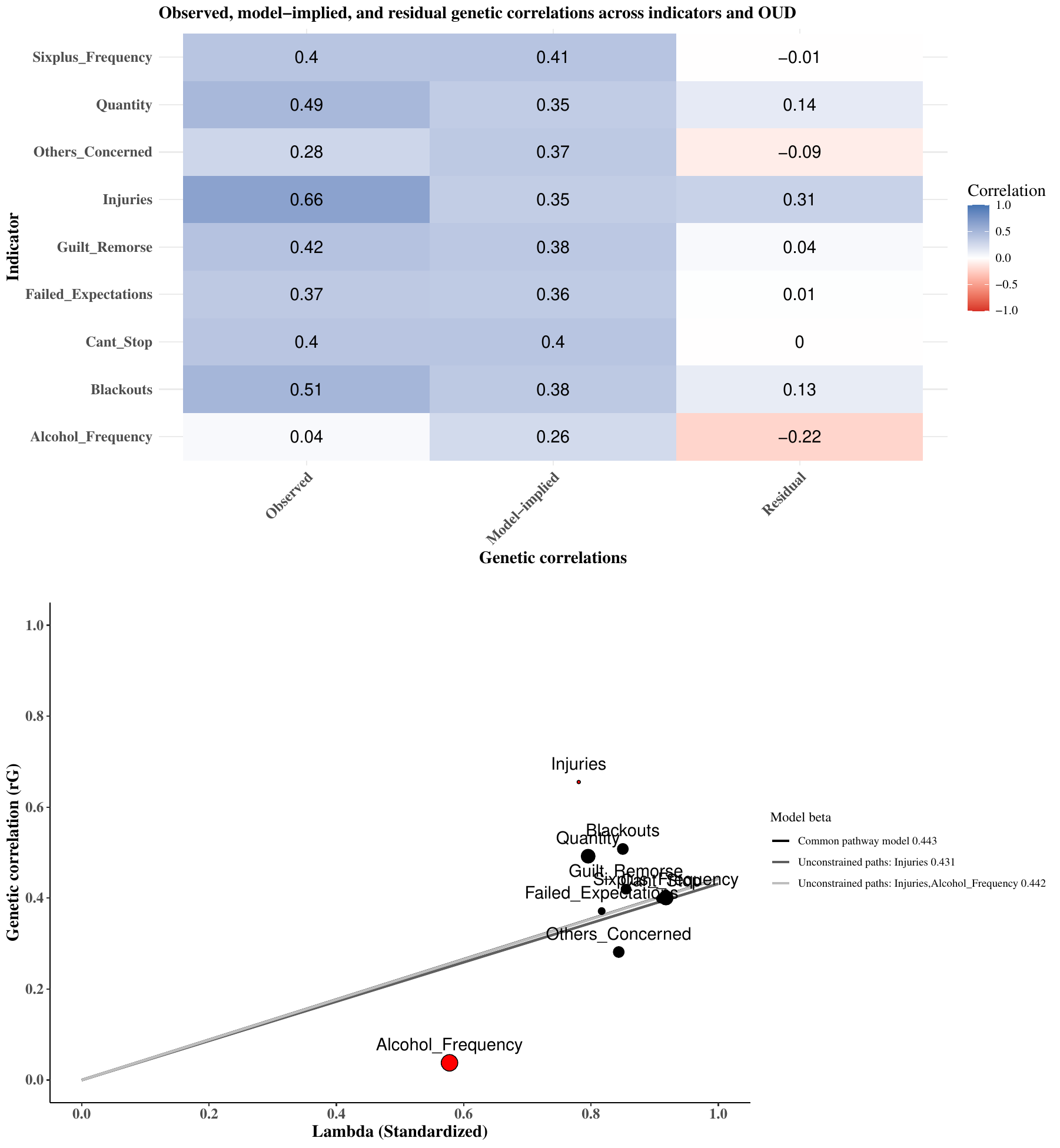

Supplementary Figure 11: Qtrait analysis of OUD

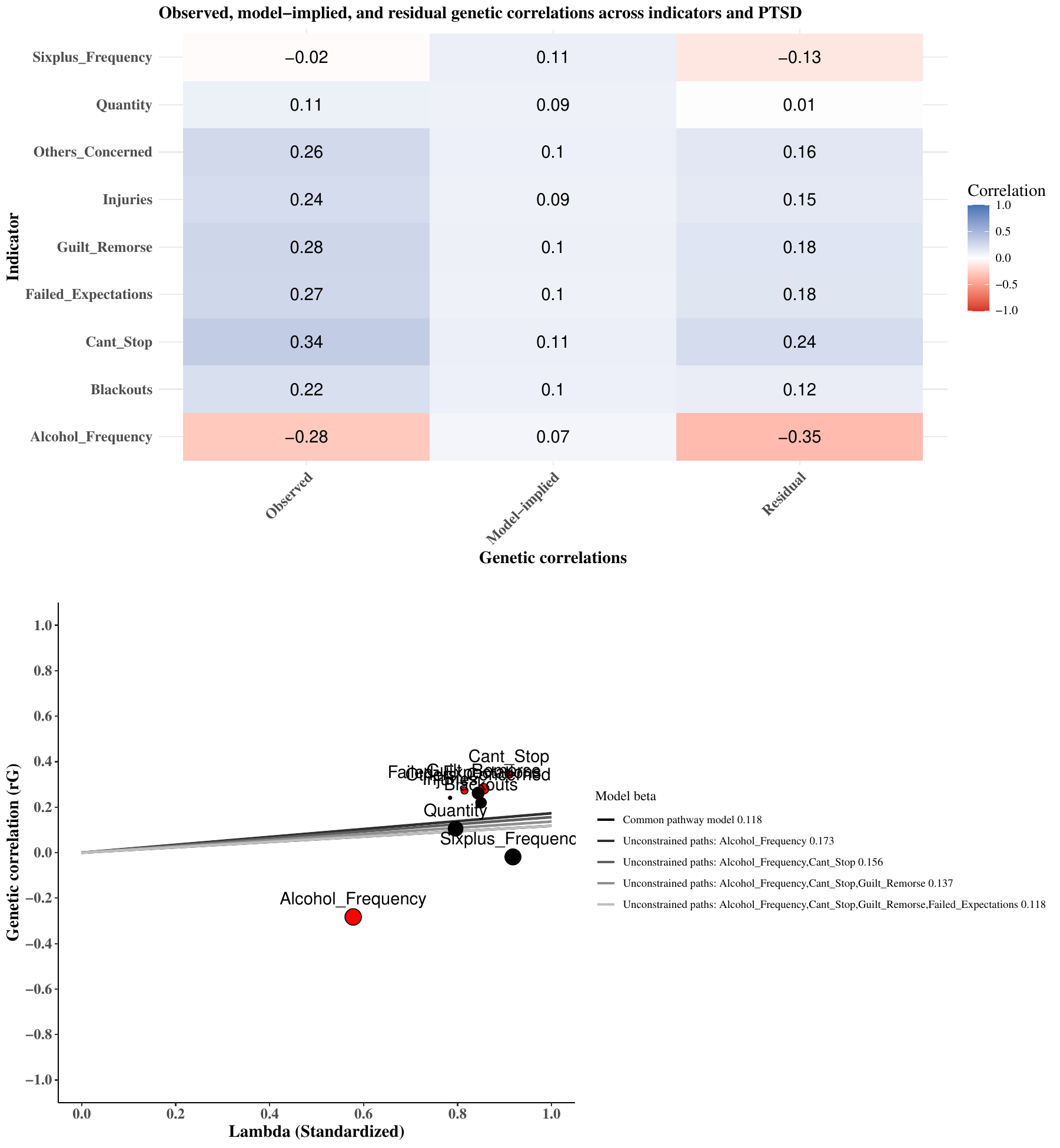

Supplementary Figure 12: Qtrait analysis of PTSD

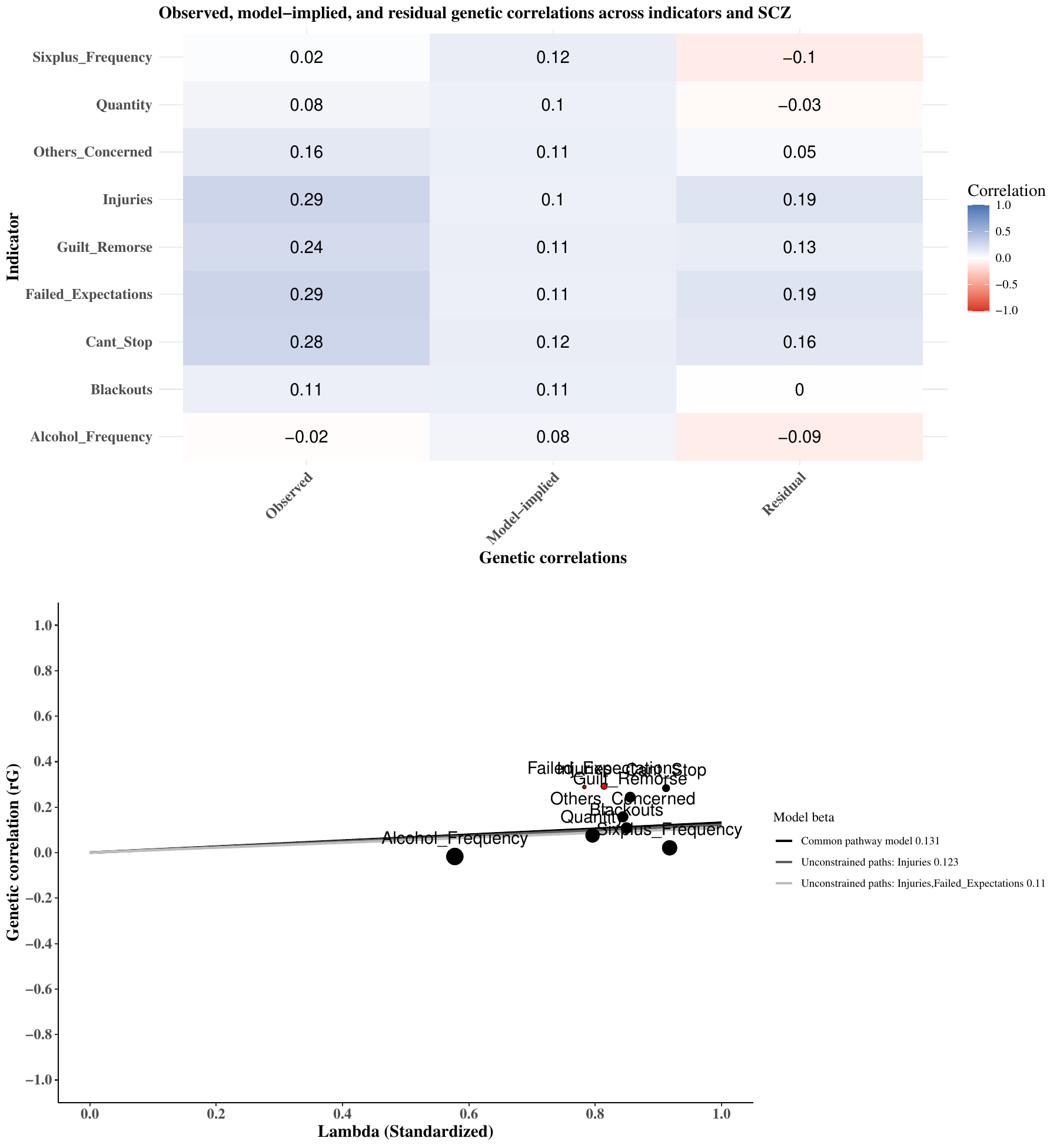

Supplementary Figure 13: Qtrait analysis of SCZ
